# The Transcriptional Landscape of Atrial Fibrillation: A Systematic Review and Meta-analysis

**DOI:** 10.1101/2023.10.30.23297750

**Authors:** Sergio Alejandro Gómez-Ochoa, Malte Möhn, Michelle Victoria Malz, Roger Ottenheijm, Jan D. Lanzer, Felix Wiedmann, Manuel Kraft, Taulant Muka, Constanze Schmidt, Marc Freichel, Rebecca T. Levinson

## Abstract

**Background:** Despite advances in understanding atrial fibrillation (AF) pathophysiology, there is limited agreement on the key genes driving its pathophysiology. To understand the genome-wide transcriptomic landscape, we performed a meta-analysis from studies reporting gene expression patterns in atrial heart tissue from patients with AF and controls in sinus rhythm (SR).

**Methods:** Bibliographic databases and data repositories were systematically searched for studies reporting gene expression patterns in atrial heart auricle tissue from patients with AF and controls in sinus rhythm. We calculated the pooled differences in individual gene expression from fourteen studies comprising 534 samples (353 AF and 181 SR) to create a consensus signature (CS), from which we identified differentially regulated pathways, estimated transcription factor activity, and evaluated its performance in classifying validation samples as AF or SR.

**Results:** Despite heterogeneity in the top differentially expressed genes across studies, the AF-CS in both chambers were robust, showing a better performance in classifying AF status than individual study signatures. Functional analysis revealed commonality in the dysregulated cellular processes between chambers, including extracellular matrix remodeling, cardiac conduction, metabolic derangements, and innate immune system activity. Finally, the AF-CS showed a good performance differentiating AF from controls in three validation datasets (two from peripheral blood and one from left ventricle samples).

**Conclusions:** Despite variability in individual studies, this meta-analysis elucidated conserved molecular pathways involved in AF pathophysiology across its phenotypes and the potential of a transcriptomic signature in identifying AF from peripheral blood samples. Our work highlights the value of integrating published transcriptomics data in AF and the need for better data deposition practices.

Graphical abstract

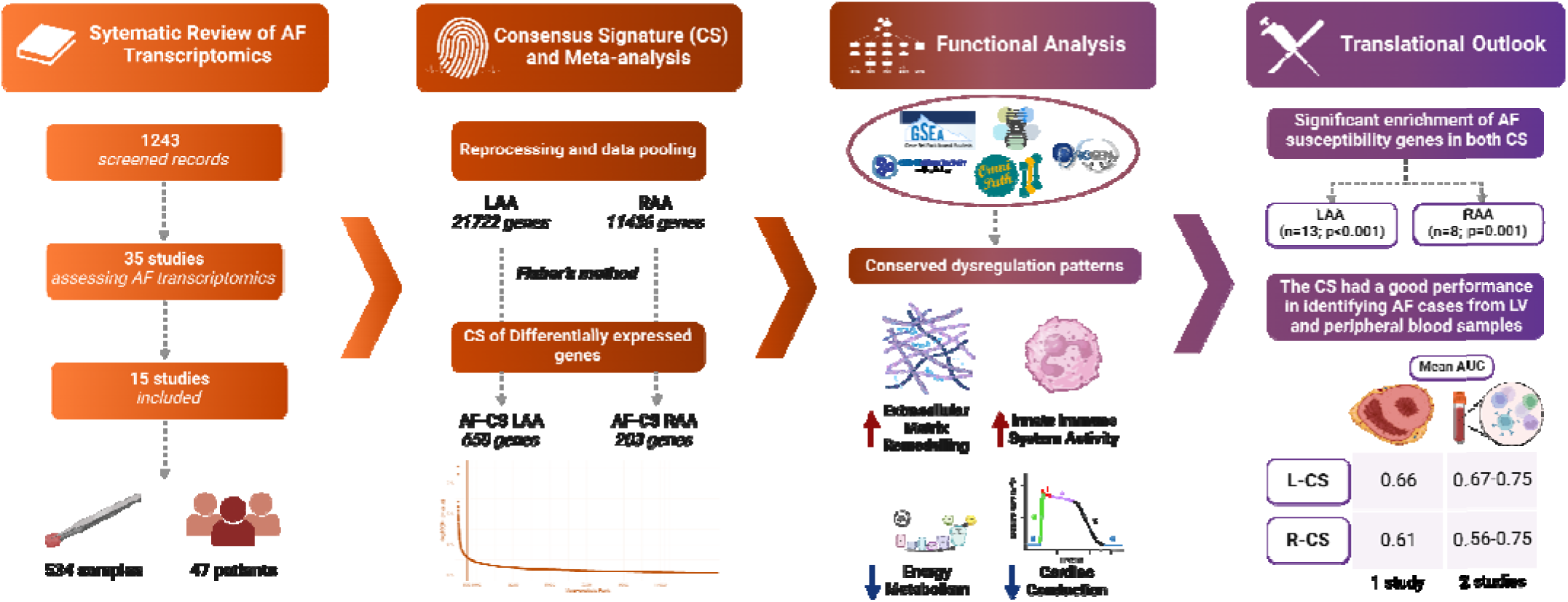

**Clinical Perspective:** *What Is New?:*

- This meta-analysis of 534 atrial tissue samples reveals a robust consensus transcriptional signature for atrial fibrillation (AF), identifying consistently dysregulated genes and pathways across heterogeneous patient populations and study designs.

*What Are the Clinical Implications?:*

- The identified consensus signature provides a molecular framework for developing targeted diagnostic biomarkers and mechanism-based treatment strategies for AF, potentially leading to more personalized and effective management approaches.
- The study’s findings of conserved gene expression patterns across different AF types and the signature’s performance in classifying AF from peripheral blood samples suggest potential for non-invasive diagnostic and prognostic applications in clinical practice.

**Research Perspective:** *What New Question Does This Study Raise?:*

- Given the identified conserved molecular pathways in atrial fibrillation (AF), how do these transcriptional signatures evolve across different AF types (paroxysmal, persistent, and permanent) and in response to various therapeutic interventions?

*What Question Should be Addressed Next?:*

- Can the consensus transcriptional signature be validated in large-scale, prospective studies to develop and evaluate blood-based biomarkers for early AF detection, risk stratification, and treatment response prediction?
- How do the identified dysregulated pathways, particularly those related to structural remodeling and metabolic reprogramming, interact with genetic predisposition and environmental factors to influence AF onset and progression?

## Introduction

Atrial fibrillation (AF) is the most prevalent sustained arrhythmia worldwide[1]. It is associated with substantial quality of life deterioration and a high risk of adverse clinical outcomes, including thromboembolic events and mortality[2]. AF is a heterogeneous disease that arises in combination with other cardiovascular diseases, such as hypertension and heart failure, and significantly contributes to chronic disease burden on the individual and the healthcare system[3, 4]. Despite the broad array of available diagnostic tools and therapeutic interventions, recurrent AF, treatment resistance, and adverse side effects of current pharmacotherapies remain pressing challenges[5]. Moreover, current therapies do not explicitly target AF’s molecular and genetic contributors, potentially limiting the possibility of optimally managing this condition[6]. Therefore, a more complete understanding of AFs underlying molecular and genetic contributors has become essential[7].

Transcriptomic analyses have emerged as a valuable tool for probing AF’s genetic and molecular foundations[8]. However, a significant gap in the existing research landscape is the lack of comprehensive studies considering the entire transcriptome[9–13]. When studies analyze the complete transcriptome, they often still focus on identifying specific genes, which may vary widely between studies, resulting in a high degree of heterogeneity in the key AF genes identified[14]. Technical differences between experiments may partially explain these differences; however, contrasts in sociodemographic and clinical features, such as age, gender, ethnicity, comorbidities, and AF type, between cohorts also represent a relevant source of variation in the observed trends[15]. Consequently, these factors may inadvertently influence the gene expression profiles associated with disease, result in patient- or population-specific genomic responses, and affect the expression levels of genes involved in the etiology of AF[16]. A consensus transcriptional signature, which could provide a more unified perspective on the molecular processes that instigate and perpetuate this disorder, could offer relevant insights into the disease and lay the groundwork for more personalized and effective treatment strategies[17].

This consensus signature meta-analytical approach using transcriptomic data has been previously applied in studies evaluating cardiac tissue from patients with heart failure (HF) and renal tissue from patients with chronic kidney disease, validating the feasibility of combining RNA-sequencing and microarray data from different sources, platforms, and conditions, as well as providing a statistical framework applicable in other contexts[18, 19]. In light of this, the primary aim of our study was to develop a consensus transcriptional signature for the left- and right atrial appendages in AF. To accomplish this, we undertook a systematic review and meta-analysis of existing studies comparing myocardial gene expression patterns in patients diagnosed with AF and control subjects, presumably in sinus rhythm (SR). Moreover, to understand the translational relevance of this signature, we aimed to assess whether this consensus signature can be leveraged in other scenarios, such as peripheral blood transcriptomics, to accurately differentiate patients with AF from controls. This consensus signature represents an open tool easily accessible by the research community with the potential of usage without significant previous data analysis experience.

## Methods

### Search strategy and selection criteria

We followed two recent guidelines for the conduct of the systematic review and meta-analysis and Preferred Reporting Items for Systematic Reviews and Meta-Analyses (PRISMA) guidelines for reporting **(Supplemental Table 1)**[20–22]. The protocol of this study is registered in the PROSPERO database with the record code CRD42023399021.

MEDLINE/Ovid, EMBASE/Ovid, CINAHL/EBSCOhost, and Google Scholar were searched to identify relevant studies from January 1st, 2000, until March 18th, 2024, without language restrictions. Search terms related to AF transcriptomics were used, including atrial fibrillation, atrial cardiomyopathy, transcriptome, microarrays, and RNA-Seq, among many others. The complete search strategy is described in **Supplemental Text 1**. All observational studies comparing the transcriptomic profile (by microarrays or RNA-Seq) of patients with a diagnosis of AF with individuals suspectedly in SR were included. Only studies that assessed differential gene expression between AF and SR groups by analysis of myocardial tissue samples and provided any type of reanalyzable data were included.

In parallel to the bibliometric database search, we queried four additional repositories that store biomedical studies datasets: the Gene Expression Omnibus database, the European Nucleotide Archive, ArrayExpress, and the European Genome Phenome Archive. The following search terms were used: “atrial fibrillation,” “atrial cardiomyopathy,” “atrial arrhythmia,” and “afib.” We included a dataset if it assessed myocardium samples, if data from at least four total patients were available, and if there was a protocol describing sample gathering and processing methodology available. **Figure 1** summarizes the main steps in the analysis process of this study[18, 19].

**Figure 1.**
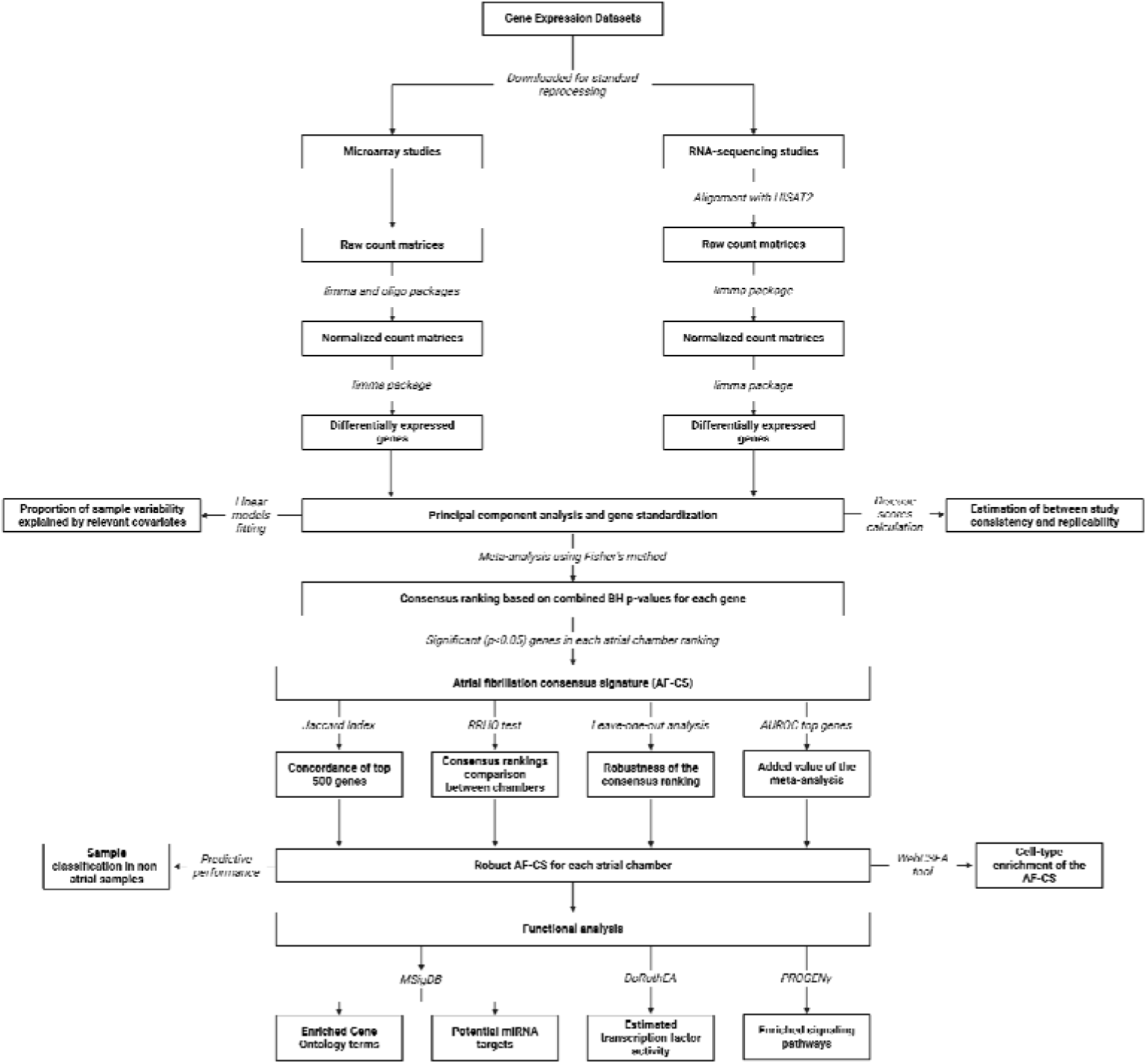
Summary of the analysis process of the atrial fibrillation consensus signature meta-analysis.

### Database screening and data extraction

Four reviewers screened titles and abstracts in duplicate to systematically identify potentially includable studies according to the selection criteria. Subsequently, four reviewers independently extracted the following data from each included study: name of the first author, the country in which the study was performed, year of publication, study design, the total number of patients, number per group (AF vs. SR), demographic characteristics of the population evaluated, type of AF evaluated, left ventricular ejection fraction, sample sources, sequencing and analysis methods, and differentially expressed genes between groups.

### Quality assessment

Four authors independently assessed each study in duplicate using an adapted version of the Checklist for Analytical Cross-Sectional Studies from the Joanna Briggs Institute (JBI) **(Supplemental Table 2)**. This adapted checklist was designed due to the lack of specific quality assessment tools for studies performing unbiased transcriptomic analysis in tissue samples and allowed an objective assessment of relevant methodological and reporting aspects of unique importance in this type of studies. It comprises ten items covering study design and participant characterization, sample processing, statistical analysis, and data reporting. Conflicts were resolved by consensus or the intervention of an additional reviewer.

### Data processing

The raw data from studies found through the systematic search strategy and from queried repositories were downloaded and processed uniformly. Non-normalized count matrices were downloaded directly, whereas, for studies with only sequencing files (.fastq files), sequence alignment was performed using the HISAT2 alignment algorithm to acquire count matrices for data derived from RNA-Seq experiments. Standard processing and normalization were performed using the limma package for datasets derived from RNA-seq, while the oligo package was also used for those derived from microarrays.

### Sample classification and contrasts

We classified the collected samples according to AF rhythm (AF vs. SR) and according to sample origin (left atrial appendage and right atrial appendage [LAA and RAA, respectively]). From this, the differential gene expression between AF and SR in each chamber was analyzed.

### Statistical analysis

Overall, a consensus transcriptional signature meta-analysis can be defined as an approach to identify consistent gene expression patterns (transcriptional signatures) across multiple independent studies focusing on a particular condition[18, 19]. This approach allows for the identification of more robust and reliable molecular signatures, potentially elucidating pivotal transcriptional programs consistently correlated with specific states or responses, such as disease onset, progression, or reaction to specific interventions. For this purpose, a Fisher’s combined probability test was used to pool the differential expression analysis results per individual genes if they were evaluated in at least 50% of the studies. For each contrast, the degrees of freedom for the statistical tests were derived from the number of studies included. Since none of the included studies mentioned having performed any probabilistic sampling procedure, additional study weighting was not performed. After the Benjamini-Hochberg (BH) correction of the pooled P values, a gene ranking representing the AF transcriptomic signature was generated[23]. Using an inferential approach, we estimated the accuracy of using individual studies to classify samples of other studies into either “AF” or “SR” categories. For this purpose, a disease score was calculated by linearly combining the t-values of the differentially expressed genes of the reference studies (disease pattern) with their expression values in the individual test study.

The number of disease classifiers built was the same as the total study number for each comparison based on the t-values of the differentially expressed genes in each contrast. Once each sample-level disease score was calculated, the area under the receiver operating characteristic curve (AUROC) was used to test the score’s accuracy to discriminate samples from AF patients, reflecting the conservation of gene regulation patterns across studies. Finally, the consistency of the direction of transcriptional regulation in the disease score results was also assessed by separating the top differentially expressed genes based on their regulation direction and performing gene set enrichment analysis (GSEA).

In addition, we assessed the concordance of the top 500 genes of each chamber’s consensus signature using the Jaccard index. Complementarily, we compared the consensus rankings of LAA and RAA using the Rank-Rank Hypergeometric Overlap (RRHO) test. This threshold-free algorithm detects common trends between two ordered lists of differentially expressed genes. In this method, the genes are ordered based on the product of the negative logarithm (base 10) of the differential expression p-value and the effect size direction[24]. According to this ranking, genes with unchanged, upstream, and downstream effects are positioned in the middle, at the top, and at the bottom of the list, respectively. The degree of similarity between the gene lists from two different datasets is then quantified through a one-sided p-value, which is determined using the principles of the hypergeometric distribution.

### Sensitivity analysis

Using a leave-one-out procedure, we evaluated how much each study impacted the overall findings. Therefore, the meta-analysis for each contrast was repeated the same number of times of the amount of studies present, each time leaving out the data from one study. After each round of analysis, we performed a Spearman rank correlation test comparing the leave-one-out result with the full meta-analysis to test the influence of the left-out study. A correlation value close to 1 would indicate that the results were almost identical, and therefore, the excluded study did not significantly impact the overall findings. Conversely, a low correlation value would highlight the studies that strongly influenced the meta-analysis. This approach allowed us to assess the robustness of the meta-analysis ranking rigorously.

### Added value of the meta-analysis

To analyze the benefit of combining the available transcriptomic data related to AF in our meta-analysis, we examined whether the top 500 genes identified from the consensus signature provided a more representative transcriptional ‘fingerprint’ of AF than the top 500 genes picked from each study. In particular, we checked whether the AUROCs based on the consensus genes were significantly better than those derived from individual studies using the Wilcoxon paired test. At first, we divided the top 500 genes into two groups: those up- and down-regulated in AF. We then checked for their presence in each individual study’s gene activity ranking using GSEA. Then, we compared these enrichment scores to the ones obtained using the top 500 genes from each separate study with the Wilcoxon paired test.

### Functional analysis

After the meta-analysis, each gene resulting −log10 p-value was weighted based on the mean direction of change across the studies to develop the directed consensus signature. From these results, gene ontology terms, miRNA targets, and pathways from the Molecular Signatures Database (MSigDB) were used to enrich the signature using the *msigdbr* package. In parallel, the activity of transcription factors (TF) was appraised using DoRothEA. Furthermore, signaling pathways activity was estimated using PROGENy. P values adjusted with the BH procedure were computed for each test.

### Tissue-cell-type enrichment of the AF-CS

Using the differentially expressed genes (BH p-value < 0.05) in each CS, we aimed to explore the potential cellular context in each chamber using WebCSEA. This tool provides a specialized query interface for analyzing gene sets against an extensive repository of cell expression profiles specific to various human tissue types. For each query run, the system produces a series of analytical metrics, including the raw statistical significance value (p-value) specific to cell type enrichment analysis (CSEA), a consolidated p-value derived using a permutation-based statistical approach, and a list of shared genes between the query set and the cell type-tissue-specific expression profiles.

### Performance of the AF-CS as predictors of AF status

Finally, we aimed to explore how the two consensus transcriptional signatures derived from atrial myocardium analyses would perform in classifying other types of samples (peripheral blood, left ventricle and epicardial adipose tissue) derived from AF and SR patients in publicly available datasets. We observed an overall good performance of both LAA and RAA AF-CS in discriminating samples from patients with AF from those from patients in SR across different tissues, especially in peripheral blood **(Supplementary Tables 4 and 5)**.

## Results

### Descriptive analysis

This systematic review and meta-analysis was performed in compliance with Preferred Reporting Items for Systematic Reviews and Meta-Analyses (PRISMA) guidelines for reporting **(Supplemental Table 1)**. Of the 1243 references screened, we identified 35 studies that applied transcriptomic approaches in atrial tissue for the study of AF[13, 25–57]. Of these, only 15 studies met the complete selection criteria by providing complete gene counts or raw sequences for each of the assessed samples, therefore being included in the meta-analysis **(Figure 2)**[25, 27, 30, 31, 33–35, 44–48, 52, 57, 58].

**Figure 2.**
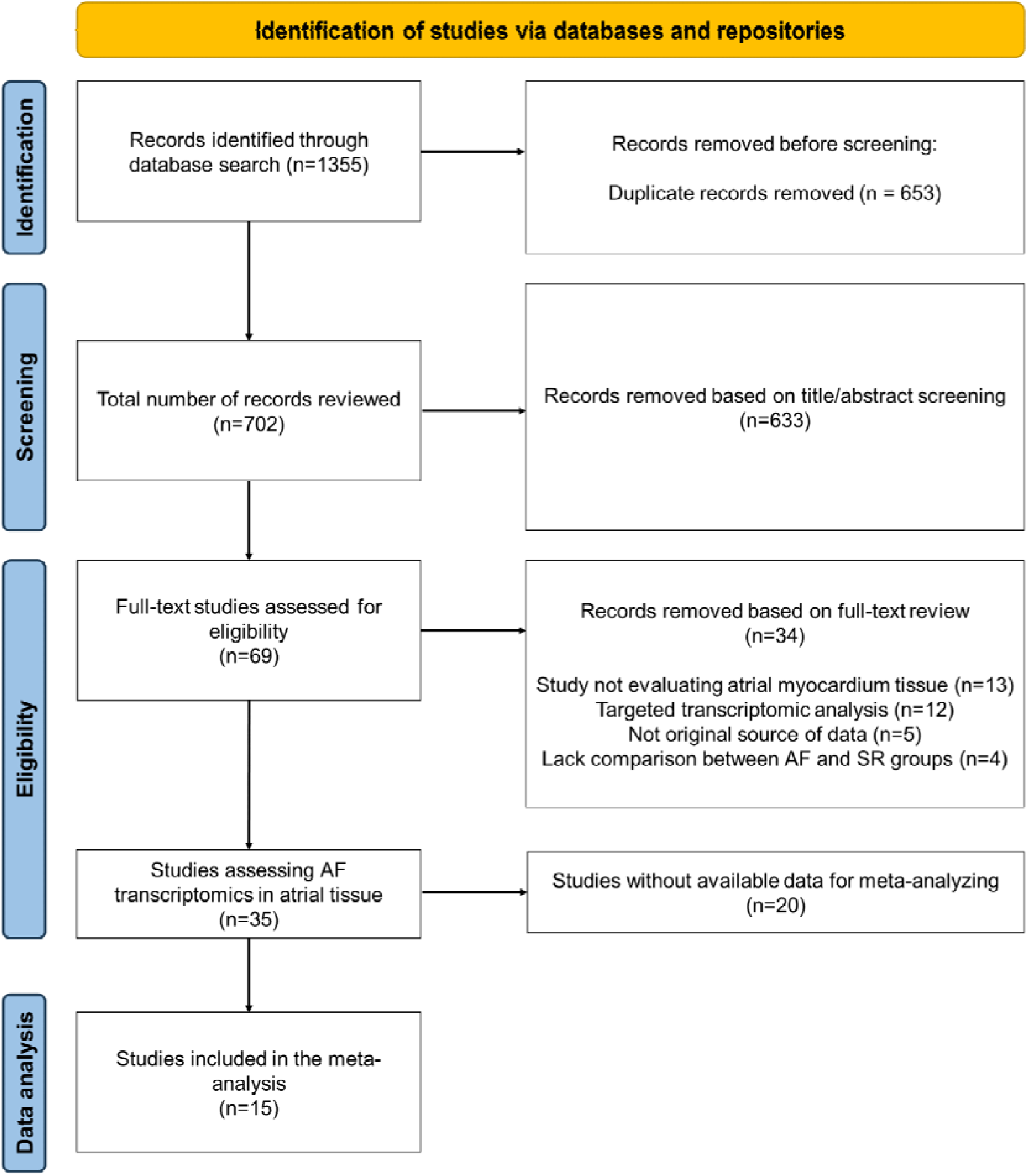
Flowchart of study selection.

The selected studies were published between 2005 and 2023, with the majority (9 [60%] of 15) published within the past five years. The data sets derived from these studies consisted of a total of 534 samples (353 AF and 181 SR) from 470 patients (mean age 58.5 years; 58% male) **(Supplemental Figure 1 and Supplemental Table 2)**[44, 59–71]. Most samples were collected in the context of valve surgery (14/15; 93%), followed by coronary artery bypass grafting (CABG) surgery (7/15; 47%) and cardiac transplantation (2/15; 13%). The majority of studies included patients with permanent or persistent AF, with only three including patients with paroxysmal AF. Nevertheless, the lack of individual-patient information limited the assessment of different AF types independently **(Table 1)**. **Figure 3** summarizes the general characteristics of the studies included in the meta-analysis, explicitly highlighting the reporting of relevant information **(Figure 3A)** and the sample sizes evaluated **(Figure 3B)**. Unfortunately, only a small minority of studies reported individual information on the included patients, which limited the performance of analyses by relevant covariates. From the total studies, 11 analyzed samples from the left atrial appendage (LAA), while eight analyzed samples from the right atrial appendage (RAA) **(Table 1)**. Finally, both microarray (n=7; 47%) and RNA-Seq (n=8; 53%) technologies were utilized **(Table 1)**.

**Figure 3.**
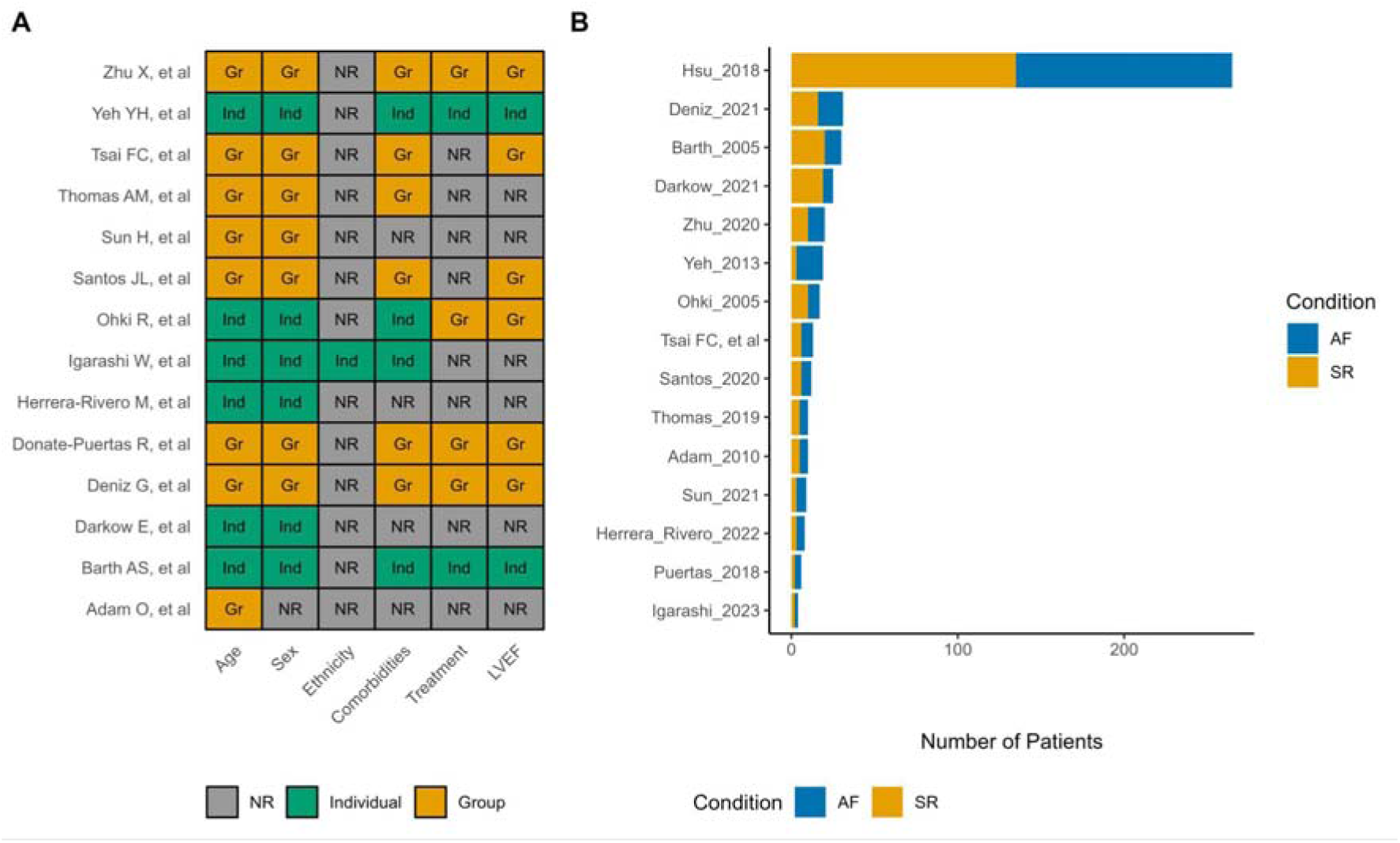
Summary of overall study characteristics. **A,** Available metadata on relevant characteristics in the included datasets. **B,** Sample size per group categories. ***Abbreviations.*** Ind: Individual-patient data; Gr: Group data (measures of central tendency and dispersion); NR: Not reported.

**Table 1.** General characteristics of the included studies.

| First Author | Year | Country | Total Sample | AF Patients | SR Patients | AF Types Evaluated | Sample Collection Context | Sample Sources | Technology | Platform |
| --- | --- | --- | --- | --- | --- | --- | --- | --- | --- | --- |
| Adam O, et al[25] | 2010 | Germany | 10 | 5 | 5 | Permanent | Valve replacement surgery | Left atrial appendage | Microarrays | Affymetrix HGU133-Plus 2.0 |
| Barth AS, et al[27] | 2005 | Germany | 30 | 10 | 20 | Permanent | Valve replacement surgery; Coronary Artery Bypass Graft surgery | Right atrial appendage | Microarrays | Affymetrix Human Genome U133A Array |
| Darkow E, et al[30] | 2021 | Germany | 25 | 6 | 19 | Permanent | Valve replacement surgery; Coronary Artery Bypass Graft surgery | Right atrial appendage | RNA-Seq | Illumina HiSeq4000 |
| Deniz G, et al[31] | 2021 | Turkey | 31 | 15 | 16 | Persistent | Valve replacement surgery | Left atrial appendage; Right atrial appendage | Microarrays | Affymetrix HGU133-Plus 2.0 |
| Doñate-Puertas R, et al[33] | 2017 | France | 6 | 4 | 2 | Paroxysmal; Persistent; Permanent | Valve replacement surgery | Left atrial appendage | Microarrays | Human Genome U133A 2.0 Array |
| Herrera-Rivero M, et al[34] | 2022 | Netherlands | 8 | 5 | 3 | Paroxysmal; Persistent | Valve replacement surgery; Coronary Artery Bypass Graft surgery; Heart transplantation | Left atrial appendage; Right atrial appendage; Ventricles | RNA-Seq | Illumina NextSeq500 |
| Hsu J, et al.[35] | 2018 | USA | 265 | 130 | 135 | Not reported | Valve replacement surgery; Coronary Artery Bypass Graft surgery | Left atrial appendage | RNA-Seq | Illumina HiSeq 2000 |
| Igarashi W, et al.[58] | 2023 | Japan | 4 | 2 | 2 | Permanent | Valve replacement surgery | Right atrial appendage | RNA-Seq | Illumina NovaSeq 6000 |
| Ohki R, et al[44] | 2005 | Japan | 17 | 7 | 10 | Not reported | Valve replacement surgery; Coronary Artery Bypass Graft surgery | Right atrial appendage | Microarrays | Human Genome U95A Set |
| Santos JL, et al[45] | 2020 | Denmark | 12 | 6 | 6 | Persistent;<br>Permanent | Valve replacement surgery | Right atrial appendage | RNA-Seq | Illumina HiSeq 2500 |
| Sun H, et al[46] | 2021 | China | 9 | 6 | 3 | Paroxysmal;<br>Persistent | Heart transplantation | Left atrial appendage | RNA-Seq | Illumina HiSeq X ten |
| Thomas A, et al[47] | 2019 | England | 10 | 5 | 5 | Permanent | Valve replacement surgery; Coronary Artery Bypass<br>Graft surgery | Left atrial appendage; Right atrial appendage | RNA-Seq | Illumina NextSeq500 |
| Tsai F, et al[48] | 2016 | Taiwan | 13 | 7 | 6 | Persistent | Valve replacement surgery; Coronary Artery Bypass<br>Graft surgery | Left atrial appendage; Right atrial appendage | Microarrays | Affymetrix HGU133-Plus<br>2.0 |
| Yeh Y, et al[52] | 2013 | Taiwan | 19 | 16 | 3 | Persistent | Valve replacement surgery | Left atrial appendage; Other | Microarrays | Human Genome U133A 2.0<br>Array |
| Zhu X, et al[57] | 2020 | China | 20 | 10 | 10 | Persistent | Valve replacement surgery | Left atrial appendage | RNA-Seq | Illumina HiSeqTM 2500 |

To identify bias in the included studies, we used a modified version of the Checklist for Analytical Cross-Sectional Studies from the Joanna Briggs Institute (JBI) which included 10 points **(Supplemental Table 3**). We observed a mean score of 7/10 points for the included studies, with no studies scoring less than four points **(Supplemental Table 3)**. The following sections describe the AF-CS results for each evaluated cardiac chamber (L-CS for the LAA and R-CS for the RAA).

### Overall characteristics and consistency measures

At first, we sought to characterize the technical characteristics of the different datasets by atrial appendage. Gene coverage analysis, which assesses the similarities between the assessed genes across studies, highlighted higher comparability of the LAA samples (mean Jaccard index of ≈0.70) than RAA samples (mean Jaccard index of ≈0.51), meaning that overall the LAA sequencing results in the different studies shared more genes between them than the RAA ones (**Supplemental Figure 2)**. Moreover, an ANOVA test revealed a significant association between the technology type ("RNA-Seq" vs. "Microarrays") and the first three PCs of the z-transformed samples (P<0.05 for the first three PCs in both atria) **(Supplemental Figure 3)**. Though, after performing read count standardization, the heterogeneity derived from technical aspects was reduced. Furthermore, we evaluated the proportion of variance explained by age, sex, and left ventricular ejection fraction (LVEF) in standardized PCA to explore the role of clinical and sociodemographic parameters in gene expression patterns. Considering that LVEF represents the most consistently reported echocardiographic parameter and has a direct relationship with atrial function, it was decided to utilize this measure in the absence of more specific parameters for atrial status. We observed that the proportion of men in the different groups was not associated with any principal component, explaining zero percent of the variance. Mean age in each group (AF and SR) explained 3.5% of the variance in LAA and 4.9% in RAA, while mean LVEF explained 4.5% in LAA and 15.3% in RAA. In comparison, the variance explained by AF diagnosis was 7.9% in the LAA and 13% in the RAA **(Supplemental Figure 4)**.

We then evaluated the consistency of the results of the studies for genes that ranked highest after differential expression analysis in each individual study **(Supplemental Figure 5)**. We compared the top 500 differentially-expressed genes (DEG) in each individual study, observing low concordance in the results of both cardiac chambers ( mean Jaccard Index LAA: 0.02 and RAA: 0.03) **(Supplemental Figure 6A and E)**. However, when evaluating the predictive capacity of these top 500 genes of each study to discriminate the diagnosis of AF in other datasets using a disease score, we observed a relatively high performance according to the median AUROC values (LAA: 0.74 and RAA: 0.78) **(Supplemental Figure 6B and 6F)**. This disease score was calculated by linearly combining the t-values of the differentially expressed genes of the reference studies (disease pattern) with their expression values in the individual test study. This was built assuming that if two or more studies share AF transcriptional signatures, the disease score derived from these could be used to discriminate between sample groups in other independent datasets accurately. Therefore, this disease score would estimate how similar the expression profile observed in each sample with the disease phenotype is, going beyond changes in the mean expression of specific genes and focusing on the joint regulation of multiple genes.

As the molecular responses showed some degree of uniformity across the evaluated studies, we proceeded further and examined whether the direction of differential regulation of the top DEG in each dataset was consistent with their observed trends in the remaining studies. For this purpose, an independent enrichment analysis was performed on the gene-level statistics derived from the top 500 up- and down-regulated genes from each study **(Supplemental Figure 6)**. This enrichment score (ES) measures how much the top DEG in one study is over-represented among the top DEG in all the other studies, providing a consistency check for the direction of gene regulation. A positive score means that a gene is often found to be overexpressed in AF across multiple studies. Conversely, a negative score would mean that a gene is often found to be less transcribed in AF samples across these studies. In our results, the median ES for the differentially up-regulated genes in the L- and R-CS were 0.38 and 0.31, respectively **(Supplemental Figure 6C and 6G)**. On the other hand, the ES values for the down-regulated genes in the L- and R-CS were −0.37 and −0.33, respectively **(Supplemental Figure 6D and 6H)**.

Moreover, a significant correlation between the AUROC values of the disease score and enrichment scores for DEGs was observed for both the L-CS (Pearson correlations of 0.77 for up-regulated genes [p<0.001] and −0.67 for down-regulated genes [p<0.001]) and the R-CS (Pearson correlations of 0.80 for up-regulated genes [p<0.001] and −0.73 for down-regulated genes [p<0.001]). This finding suggests that, despite study-specific variations, the direction of gene expression changes was consistent across studies. Additional analyses showed similar results when different numbers of top genes were selected (50, 100, 200, 500, and 1000) **(Supplemental Figure 7)**.

### Meta-analysis of transcriptional signatures in AF

After confirming the feasibility of integrating the assessed data, we developed the AF-CS for each atrial chamber by pooling the individual-study results of 21,722 genes for the LAA and 11,436 for the RAA contrasts using the Fisher’s Method **(Supplemental Table 4)**. This consensus ranking represents the overall differential gene regulation patterns across the AF spectrum, highlighting genes consistently deregulated in common directions over multiple studies **(Supplemental Figure 8)**. In the LAA, 650 genes showed a BH p-value < 0.05; this number was 203 for the RAA analysis **(Supplemental Table 4)**. Several genes related to structural cardiac remodeling processes were observed among the top positions of the AF-CS in both cardiac chambers, including COLQ, ANGPTL2, LBH, and COL21A1[72–75]. Other genes near the top of the consensus included those involved in myocardial electrical remodeling processes, such as CACNA1G, HCN4, and KCNK3[76–78]. Multiple genes related to the activity of the innate immune system were also found to be significant and highly ranked in the AF-CS, especially in the LAA, with ASAH1, CKAP4, STING1, and FCER1G standing out. Finally, new genes not previously related to AF in the literature were identified as occupying leading positions in the consensus, including DHRS9, CRTAC1, RASL11B, and GRIP2 **(Supplemental Figure 9)**.

The leave-one-out sensitivity analysis confirmed the robustness of the overall ranking results, as the resulting gene ranks were consistently preserved after removing one study at a time (mean Spearman rank correlation: LAA=0.96 and RAA=0.94) **(Supplemental Figure 10)**. There were no significant associations between the sample size of each analyzed study and the enrichment of its specific DEG in the higher positions of the CS (LAA: Spearman correlation, 0.58, P=0.07; RAA: Spearman correlation, 0.63, P=0.15). Despite the small sample sizes observed in some studies, this result indicated that most studies were robust enough to provide real biological insights from the contrasts performed.

When analyzing the top 500 genes of each individual chamber’s consensus signature, we observed a low concordance of the genes occupying these positions (Jaccard index value: 0.17). However, a threshold-free approach (RRHO test) of the whole gene rankings revealed a significant overlap in both directions of gene expression between the two chambers, highlighting a robust overall concordance between the two signatures **(Supplemental Figure 11)**.

We then assessed the added value of performing a transcriptional consensus meta-analysis in this context by comparing the performance of the disease score constructed using the top 500 meta-analysis genes vs. individual studies. We observed superior predictive performance of the measures derived from the meta-analysis in both LAA (mean AUC AF-CS: 0.82 vs. mean AUC individual studies: 0.74. p<0.001) and RAA (mean AUC AF-CS: 0.90 vs. mean AUC individual studies: 0.77. p<0.001). Similarly, the mean enrichment scores obtained from the AF-CS results were significantly better compared to the ones from individual studies for both up-regulated genes (LAA: mean ES 0.52 vs. 0.38. p<0.001; RAA: mean ES 0.47 vs. 0.30. p<0.001) and down-regulated genes (LAA: mean ES −0.42 vs. −0.33. p=0.002; RAA: mean ES −0.50 vs. −0.37. p<0.001). When testing whether the top meta-analysis genes of each chamber’s AF-CS could predict the disease condition in samples from the other chamber, we observed significantly better performance in terms of the median AUC compared with the pairwise classifier from individual study data (LAA CS in RAA samples: 0.89 vs. individual RAA studies: 0.77. p<0.001; RAA CS in LAA samples: 0.77 vs. individual LAA studies: 0.74. p<0.001). These results suggested more consistent biological signals with the meta-analysis results over individual analyses and supported the presence of common gene expression patterns in both chambers.

### Functional analysis of the AF-CS

The AF-CS results allowed us to characterize the cellular processes consistently deregulated in AF. To this end, we estimated the activity of transcription factors, signaling pathways, and miRNAs in each of the cardiac chambers using a gene set enrichment analysis approach **(Figure 4)**. We found significant enrichment for the LAA consensus signature for 366 Gene Ontology terms, 28 Hallmark gene sets, 42 Reactome pathways, and 25 KEGG pathways **(Supplemental Table 5)**. Positively enriched gene sets in this chamber were predominantly related to epithelial-mesenchymal transition, extracellular collagen matrix organization, and cytoskeletal regulation. In contrast, negatively enriched sets were associated with diverse processes related to electrical/muscle function, neuronal signaling, muscle contraction, and ion channel complexes. On the other hand, the number of results in the R-CS enrichment was lower, with only 15 significantly enriched Gene Ontology terms. These included upregulated processes related to extracellular collagen matrix organization; however, most of the enriched terms were observed to be downregulated in the AF group, highlighting cardiac conduction, autonomic function, action potential, and cell-cell signaling processes. When comparing the top results between the chambers, collagen extracellular matrix organization processes were significantly upregulated in both atrial appendages during AF, underlining the role of extracellular matrix remodeling in AF pathophysiology. On the other hand, processes related to normal cardiac conduction and signaling mechanisms were universally downregulated in AF. Notably, we did not find processes that were significantly upregulated in LAA and simultaneously downregulated in RAA, or vice versa, suggesting that differential regulation of processes between atrial appendages, if present, is likely to be nuanced.

**Figure 4.**
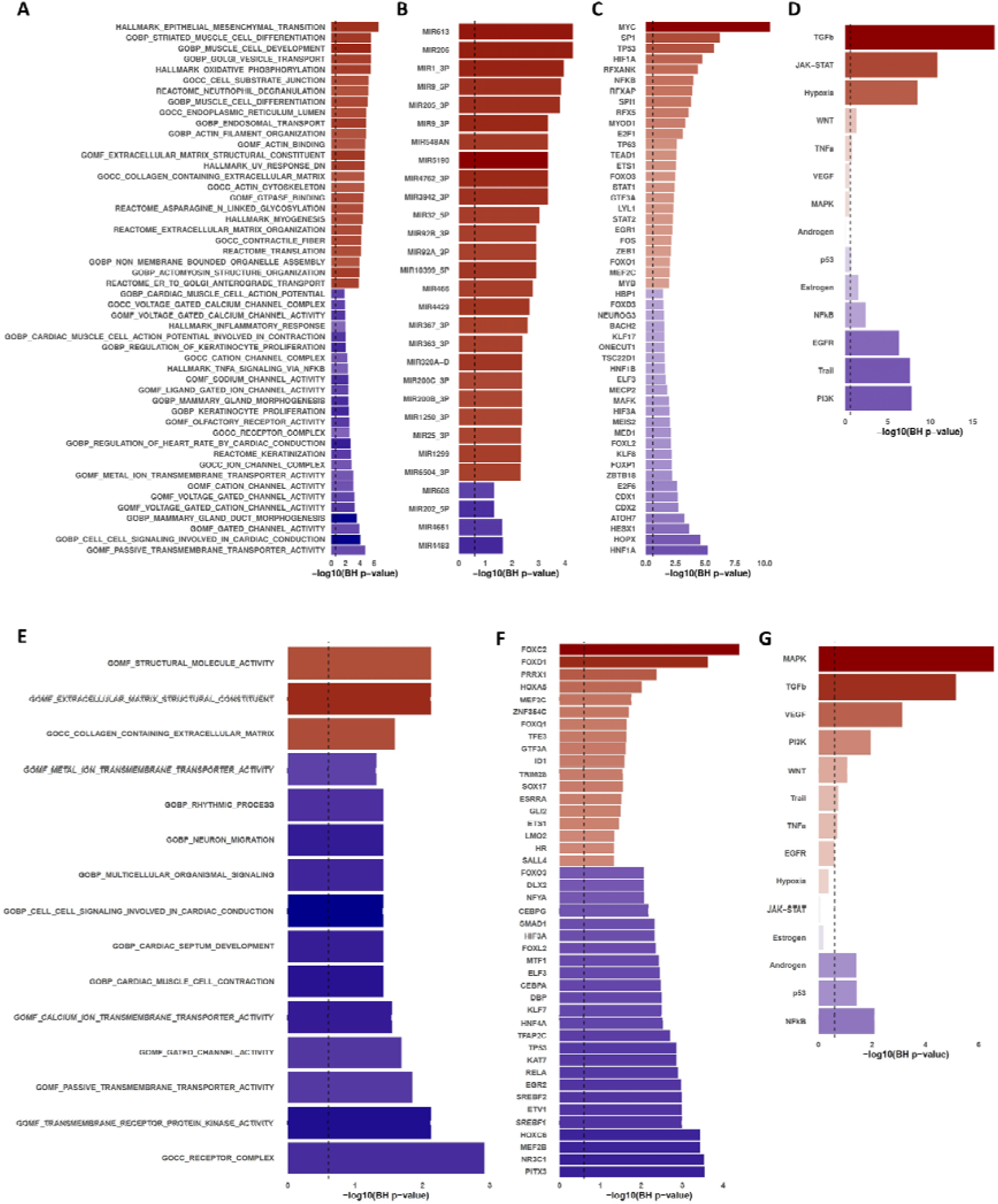
Functional analysis results of the AF-CS in each cardiac chamber.

Furthermore, the inferred transcriptional activity of 754 transcription factors (TF) allowed us to identify 100 differentially active TF in the LAA and 74 in the RAA. At the same time, 100 miRNA targets were differentially enriched in AF patients in the LAA **(Supplemental Table 2)**. Finally, pathway analysis reported the up-regulation of the TGFβ, JAK-STAT, Hypoxia, and WNT pathways in the LAA, with a simultaneous down-regulation of the PI3K, TRAIL, EGFR, NF-kB, and Estrogen pathways in this chamber. In contrast, the RAA results suggested an up-regulation of the TGFβ, MAPK, PI3K and VEGF pathways, while the Androgen, p53 and NF-kB pathways were observed to be downregulated. These results highlighted the divergent transcriptional landscapes and the potential chamber-specific molecular changes that underlie the complexity of atrial fibrillation pathophysiology **(Supplemental Table 2)**.

Bar length indicates the statistical significance of the specific item (gene set, miRNA, TF, pathway) enrichment (−log10 BH P-values) with the vertical dashed line indicating the statistical significance threshold (Benjamini-Hochberg P=0.25) and bar color indicating the direction of enrichment (A, B, E, and F) or the direction of activation (C, D, G, and H). Top 25 (A) most enriched canonical, ontology, and hallmark gene sets, along with Reactome and KEGG pathways, (B) miRNAs’ targets, (C) transcription factor activities, and (D) signaling pathway activities in each direction from the L-CS. Top 25 (E) most enriched canonical, ontology, and hallmark gene sets, along with Reactome and KEGG pathways, (F) transcription factor activities, and (G) all signaling pathway activities in each direction from the RAA AF-CS.

### Tissue-cell-type enrichment of the AF-CS

To get insights into how processes enriched in the AF-CS may relate to the function of individual cell types, we performed a tissue- and cell-type-specific enrichment analysis using WebCSEA (see Methods). At the selected Bonferroni-corrected significance threshold (p = 3.69 x 10^-5), the signatures of two sets of fetal cardiomyocytes were significantly enriched in the L-CS (Fetal_HCA_Heart_Cardiomyocytes: p-value= 8.43 x 10^-6 and FetalHeart_Ventricle cardiomyocyte: p-value= 2.63 x 10^-5), while the signatures of a fibroblast cell-type (AdultArtery_Fibroblast: p-value= 1.61 x 10^-5) and a fetal mesenchymal progenitor (FetalBrain_Fetal mesenchymal progenitor: p-value= 2.67 x 10^-5) were significantly enriched in the RAA CS **(Supplemental Figure 12)**. These results highlight the reactivation of fetal cellular programs during the cardiac remodeling process in AF-affected atria.

### Enrichment of GWAS-derived AF genes in the AF-CS

We explored whether the AF consensus signature included genes previously observed to be relevant for AF susceptibility in GWAS studies. To this end, we tested whether the number of AF susceptibility genes by GWAS in the NHGRI-EBI GWAS Catalog was significantly higher in the consensus AF transcriptomic signature of each chamber than would be expected by chance, observing a significant enrichment of AF susceptibility genes in both the LAA (n=13; p<0.001) and RAA (n=8; p=0.001) consensus signatures[79]. Among the shared genes we highlight the interleukin-6 (IL-6) receptor (IL6R), as single nucleotide variants (SNV) at this gene locus have been associated with AF development and recurrence after catheter ablation and its expression was consistently dysregulated in our AF consensus signatures for both chambers[80–84].

### Performance of the AF-CS as predictors of AF status

We evaluated the predictive potential of the AF-CS derived from atrial myocardium in two distinct contexts: peripheral blood and left ventricular myocardium derived from AF and SR patients in publicly available datasets[85–87]. This approach aimed to assess the broader applicability and potential clinical utility of these signatures.

First, we explored the performance of AF-CS in peripheral blood samples, considering its potential as a non-invasive biomarker for AF. For this purpose, we tested different numbers of top genes from the consensus regarding their performance in sample classification to find the optimal count for each chamber’s signature. As a results, utilizing the top 300 genes from the L-CS and the top 200 genes from the RAA AF-CS, we observed promising discriminatory power between AF and SR patients (AUROC: 0.66 - 0.75 for LAA; 0.56 - 0.75 for RAA).

We then extended our analysis to left ventricular myocardium samples from patients with nonischemic (NICM; n=8) and ischemic (ICM; n=8) heart failure reported in *Yang et al*[85]. f. Interestingly, the AF-CS maintained discriminatory power in this context, suggesting a broader cardiac remodeling process in AF that extends beyond the atria.

These findings highlight the robustness of our AF-CS and its potential translational value, particularly in developing blood-based biomarkers for AF detection and monitoring. Detailed performance metrics for each tissue type and dataset are presented in **Supplemental Table 6** (L-CS) and **Supplemental Table 7** (R-CS), highlighting a significantly better performance of both CS for identifying AF cases from LV samples from the study of *Yang et al* compared to individual studies. On the other hand, only the L-CS had a superior performance than individual studies in classifying samples to AF or SR status from peripheral blood samples.

## Discussion

The present transcriptional meta-analysis included 534 samples from 470 patients in 15 studies, representing the largest meta-analysis of transcriptomic data in AF performed to date. Collecting and analyzing this data presented significant challenges, primarily due to the heterogeneity of the original studies in terms of patient populations, sample collection methods, and data processing techniques. Considerable effort was invested in standardizing the data across studies, including rigorous quality control measures, batch effect correction, and the use of a disease scoring metric to enable cross-study gene expression comparability. The robust consensus gene rankings revealed a consistent dysregulation in genes involved in structural and electrical cardiac remodeling, also uncovering novel genes not previously associated with AF. Functional analysis delineated consistently deregulated cellular processes such as epithelial-mesenchymal transition and collagen extracellular matrix organization, while tissue-cell-type enrichment analysis highlighted a significant enrichment of fetal cardiomyocytes in the LAA and fibroblast and fetal mesenchymal progenitor cells in the RAA. Furthermore, we observed a significant enrichment of AF susceptibility genes reported in previous GWAS studies in both consensus signatures. Finally, the AF-CS had a better performance than individual studies in the classification of non-atrial samples according to the AF status. Despite the methodological challenges of meta-analyzing datasets with limited meta-data, our work highlights the value of integrating heterogeneous transcriptomic datasets to identify conserved molecular pathways, thereby contributing to our understanding of AF pathophysiology.

Considering the complex pathophysiological landscape of AF, identifying conserved molecular mechanisms and cellular responses from a transcriptomics perspective represents a significant opportunity to improve our understanding of the phenomena underlying this arrhythmia[88]. Overall, the main processes highlighted in the consensus were related to the structural remodeling of the myocardium[19]. In particular, the functional analysis highlighted the relevance of epithelial-mesenchymal transition (EMT), a widely studied process in HF[89–91]. EMT is characterized by the reactivation of a fetal cellular program involving the expression of embryonic markers in response to myocardial injury[89]. This phenomenon was also observed in the results of the cell type-specific enrichment analysis, suggesting the emergence of cells with a fetal-like phenotype in the AF atria, at least from a genomic perspective. Furthermore, epicardial EMT in response to injury is also associated with increased fibroblast activation and their transformation into myofibroblasts under pathological stress, a finding consistent with the observed enrichment of extracellular matrix-related processes and the fibroblast cell type[89]. The resulting structural alterations and fibrosis development leads to conduction disorders and the reentry phenomena characteristic of AF[88].

Electrophysiological remodeling and calcium (Ca^2+^) handling anomalies were also consistently observed in AF tissues, underlining significant dysregulation of key potassium and calcium channels. These findings were reflected in the significant enrichment of processes pertaining to the action potential, voltage-gated channel activity, and cardiac conduction, all of which were among the top downregulated pathways in both atria. Prior studies have characterized the electrical remodeling present in AF, highlighting the relevance of action potential duration (APD) heterogeneity in the initiation and persistence of reentrant arrhythmias in AF[92, 93]. Concurrently, Ca^2+^-handling abnormalities, highlighting RyR2 dysfunction, altered L-type Ca^2+^ currents, and increased spontaneous Ca^2+^ release from the sarcoplasmic reticulum have been implicated as key drivers of AF[94]. Our results are in line with these findings, revealing a conserved dysregulation in critical calcium channels and associated proteins (S100A8, S100A9, SMOC-2) in AF atria, which may support the synergistic role of abnormal Ca^2+^ signaling and structural remodeling in failing cardiomyocyte adaptation to abnormal calcium influx and overload derived from rapid atrial stimulation, thereby facilitating the arrhythmia onset and perpetuation[95].

Furthermore, our findings suggest the presence of metabolic reprogramming processes in AF. These are closely related to EMT, as studies performed in failing hearts have shown that the fetal phenotype is characterized by a “switch back” to glycolysis as the primary energy source of the cardiomyocytes[96]. However, this change may not be straightforward, as the results of our functional analysis suggested an increase in both the activity of oxidative phosphorylation and glycolysis pathways. This apparently discordant finding is consistent with the results of two proteomic studies that reported the upregulation of proteins involved in fatty acid uptake into the mitochondria and β-oxidation, such as fatty acid translocase (FAT/CD36), (14)C-palmitic acid and CD36, which resulted in an apparent increased activity of the oxidative phosphorylation pathway in the AF groups[97, 98]. Conversely, they also demonstrated decreased expression and activity in proteins pivotal to oxidative phosphorylation, the TCA cycle, and, notably, the electron transport chain[97, 98]. This dichotomy underlines a potentially maladaptive metabolic response to the chronic energy stress inherent in AF, emphasizing that cellular efforts to augment energy production via selective protein upregulation are insufficient to offset the mitochondrial derangements in ATP synthesis[99, 100].

The present consensus signature analysis also provided relevant insights regarding the role of the innate immune system in AF, highlighting neutrophil degranulation as the top-ranking Reactome pathway in the L-CS. This finding is supported by current evidence published in the literature, which highlights increased neutrophil extravasation in the atria of AF patients, in addition to observing significant colocalization between enzymes such as myeloperoxidase and areas of atrial myocardial fibrosis[101–103]. On the other hand, the neutrophil-lymphocyte ratio has been shown to perform favorably in predicting de novo AF, as well as relevant clinical outcomes such as stroke[104–106]. Our results extend these findings across the spectrum of AF, highlighting the activity of the innate immune system as a preserved process relevant in the development and persistence of this arrhythmia.

### Strengths and limitations

As a meta-analysis with a standard bioinformatics analysis process, our study was strengthened by the increased sample size compared to individual studies, as well as the feasibility of pooling heterogeneous data, taking into account relevant factors such as the sampling/technical variability and variability derived from significant demographic and clinical covariates. Furthermore, our disease scoring metric enabled cross-study gene expression comparability. Significant directional consistency across studies supported the robustness of our differential gene regulation findings. Notably, including both atrial chambers in our meta-analytic framework provided a granular, chamber-specific perspective on AF pathophysiology, while comprehensive functional analyses yielded actionable insights into the implicated molecular processes. Finally, the validation of the consensus performance in classifying AF cases from other types of samples, specially blood samples, highlights not only the consistency of the signal across tissues and sources, but the potential of transcriptomics-derived markers for future applications in the clinical setting.

Notwithstanding its strengths, our study is subject to several limitations. First, the lack of individual patient data in most incorporated studies limited the possibility for subgroup analysis, and precluded optimal covariate adjustment. Additionally, the studies that met our inclusion criteria included predominantly patients with persistent or permanent AF. Moreover, the low proportion of dual-chamber samples and disparities in gene coverage and sample size between the LAA and RAA impede direct chamber-to-chamber comparability. Another limitation relates to the meta-analysis process itself, as our gene standardization approach could only partially eliminate technical heterogeneity arising from using varied platforms.

## Implications for Clinical Practice and Future Directions

The consensus transcriptional signatures derived from this meta-analysis, which consolidates the available knowledge from genome-wide transcriptomics, offer an opportunity to refine the current clinical approach to AF. Identifying consistently dysregulated genes and pathways across heterogeneous patient samples may represent the first step towards identifying and validating novel targeted diagnostic/prognostic biomarkers and provides a molecular framework for designing mechanism-based treatment strategies.

Despite these advances, it is paramount to address the current study’s limitations to foster future research directions. The scarcity of individual patient-level data necessitates further studies to confirm these findings in diverse patient populations, focusing on subgroup analyses to determine the potential effect of concomitant comorbid conditions and to characterize the conserved gene expression patterns in AF across the different types (paroxysmal, persistent, and permanent) to better understand the mechanisms potentially associated with arrhythmia progression and perpetuation. Finally, while we believe our study marks a significant step forward in our understanding of AF, it also delineates the critical avenues for future research aiming to translate these molecular insights into actionable clinical interventions. The stable reference provided by our consensus signatures can serve as a foundation for developing and validating targeted therapies, as well as for identifying novel biomarkers for early detection and risk stratification in AF. This underscores the importance of our work not only in advancing our understanding of AF pathophysiology but also in paving the way for more personalized and effective management strategies for this complex arrhythmia.

## Acknowledgments

The authors thank Ricardo O. Ramírez Flores for the valuable feedback and critical assessment of the manuscript.

## Data Availability

The study data are available in the supplementary material. The code used to generate the results and the processed data is available in GitHub (https://github.com/thelevinsonlab/AF_CS_MA).

## Ethical Approval

Ethical approval was not required.

## Registration Number

The study was registered in the International Prospective Register of Systematic Reviews (PROSPERO) under the record code CRD42023399021. Available from: https://www.crd.york.ac.uk/prospero/display_record.php?ID= CRD42023399021.

